# Conceptualizing and Validating a Health Worker Mobility Score to Strengthen Retention in Nigeria’s Health System

**DOI:** 10.64898/2025.12.23.25342945

**Authors:** Chukwuemeka Azubuike, Yusuf Ajiboye, Onyema Ajuebor, Oyebanji Filani

## Abstract

Nigeria faces an escalating crisis of health worker attrition and international migration, driven by persistent push factors such as low remuneration, poor working conditions, insecurity and limited career progression. Existing human resources for health (HRH) information systems are largely retrospective and descriptive, providing insufficient visibility into the behavioral signals that precede internal mobility or international migration. This paper conceptualizes and proposes the validation of a Health Worker Mobility Score (HWMS)—a behaviour-based, real-time predictive analytics tool designed to identify early warning signs of attrition among Nigeria’s health workforce.

We developed a composite, weighted mobility score informed by labour-market theory, migration theory, and human capital theory. Eight behavioural indicators—captured routinely through ManagedMedics, a novel digital HR management platform—form the score’s core domains, including job stability, locum intensity, absenteeism, training participation and digital engagement. We propose a prospective 18-month longitudinal cohort study across six Nigerian states (Lagos, Federal Capital Territory, Kaduna, Rivers, Imo, Ekiti), recruiting 800–900 health workers from approximately 200 private hospitals. Predictor data will derive exclusively from ManagedMedics platform logs, while administrative HR records and periodic surveys will serve as validation data. Predictive accuracy will be assessed using logistic regression, Cox proportional hazards models, and machine-learning classifiers (random forests, XGBoost, SVMs), following TRIPOD guidelines.

The HWMS provides a structured mechanism for transforming micro-level behavioural signals into attrition-risk tiers (low, moderate, high). Conceptual illustration shows how diverse behavioural indicators aggregate into meaningful predictors of internal mobility and potential migration. Proposed use cases span early-warning analytics, targeted retention strategies, state-level and facility-targeted workforce planning, integration with national HRH systems, and operationalization of Nigeria’s managed migration policy.

The HWMS represents an innovative, context-appropriate approach to strengthening HRH stability in Nigeria. By shifting from retrospective enumeration to predictive intelligence, the score offers policymakers, hospital managers and development partners a tool for proactive retention, ethical recruitment, and data-informed workforce planning. Future empirical validation will determine the score’s predictive accuracy, refine indicator weights and assess its feasibility for national scale-up and integration into Nigeria’s digital health ecosystem.

## 1. Introduction

Nigeria’s health system faces a chronic and multifaceted crisis of human resources for health (HRH), threatening its ability to deliver quality care and achieve universal health coverage (UHC). Despite several decades of reforms, the country continues to grapple with an insufficient, unevenly distributed, and poorly retained health workforce. With only 18.3 skilled health workers per 10,000 population, Nigeria falls significantly below the World Health Organization’s (WHO) contextual threshold of 44.5 per 10,000^1^, and is thus listed among the 55 countries on the WHO’s Health Workforce Support and Safeguards List: a signal of its acute vulnerability to health worker shortages and external migration pressures.

At the heart of this crisis lies a dangerous confluence of inadequate workforce production and distribution, inability to absorb unemployed health workers, and poor conditions of service leading to high attrition rates^5^. On the one hand, Nigeria’s health training institutions produce approximately 5,000 doctors annually, yet about 40% emigrate within the same year, mostly at the end stage of a cascade of mobility events that begin with job-hopping and internal migration within the country^2^. On the other hand, a 2022 study by Salau et al. found that 56% of workers providing direct care interventions in private hospitals in southwestern Nigeria had clear intentions to leave their roles within one year^3^, even before factoring in foreign migration: a harbinger of the endemic instability in the sector.

Health worker mobility, especially among doctors, nurses, and midwives, has become a core challenge to health service delivery in Nigeria, contributing to frequent service disruptions, inconsistent care quality, and increased institutional costs due to staff turnover. Two intertwined forms of mobility drive attrition. Internal mobility refers to health workers switching between facilities, sectors or geographic areas; this often results in maldistribution, with rural facilities severely understaffed. Out-migration or international migration involves health workers leaving Nigeria, predominantly for the United Kingdom, Canada and the United States. Key mobility drivers include: low and irregular salaries, limited career progression, poor working conditions, insecurity, and social considerations. These factors form a continuum, with behaviors such as job switching, absenteeism, and credential verification often signaling imminent exit.

Notably, many of the health workers who eventually migrate internationally begin their journey with internal job changes motivated by dissatisfaction, poor remuneration, or a desire to fund licensing exams and relocation plans. Yet, existing HRH data systems in Nigeria such as the National Human Resources for Health registry deployed on the Integrated Human Resources for Health (IHRIS) platform, are largely retrospective, focusing on enumeration credentialing and training, with limited implemented capacity to anticipate future health workforce movements or risks.

In response, the National Policy on Health Workforce Migration (2023) highlighted the urgent need to institutionalize managed migration and emphasizes improving retention rates, promoting ethical recruitment, enhancing data collection for real-time workforce planning^4^, however no real effect has been seen on health worker migration patterns. Other national policy responses, such as the 2020 National Human Resources for Health Policy and accompanying 2021–2025 Strategic Plan, have sought to address structural HRH constraints through improved planning and production, they have yet to meaningfully address the health worker migration issue or offer frameworks for early intervention^6^.

The report of the 2023 HRH reform assessment commissioned by the Nigeria Health Commissioners Forum (NHCF) and USAID’s Health Workforce Management (HWM) Activity corroborates this gap, noting that HRH management efforts across states remain modest, fragmented, and reactive. The report also highlighted a weak culture of proactive workforce planning and inadequate use of digital tools for real-time workforce analytics.**^Error! Bookmark not defined.^**

## 2. Literature Review

Organizations today are increasingly turning to predictive analytics to revolutionize their HR decision-making processes^5^. By leveraging the power of artificial intelligence and data-driven insights, HR professionals can gain valuable predictive capabilities to anticipate future talent needs, identify high-performing candidates, and improve employee retention rates^6^. Recent advances in predictive workforce analytics demonstrate that machine-learning models can meaningfully forecast nurse turnover and turnover intention across diverse health systems. Evidence from Korea, for example, shows that decision-tree and random-forest models can predict actual turnover with very high accuracy using indicators such as salary, age, and employment status^7^.

In the United States, Naïve Bayes classifiers applied to electronic medical record audit-trail data have successfully identified behavioural precursors to exit, including documentation patterns and workload signals^8^. Studies of turnover intention further reinforce the relevance of psychosocial and behavioural predictors: logistic-regression models in South Korea highlight job stress and sleep disturbance as strong determinants of intention to leave^9^. Together, these studies show that predictive modelling, whether based on administrative, behavioural, or psychosocial markers, offers actionable insight for workforce planning.

In low- and middle-income countries (LMICs), HRH analytics have primarily focused on enumeration, credentialing, distribution mapping, training etc. Tools such as the open source integrated Human Resources Information System (**iHRIS)** – which is used in Kenya, Uganda, Zambia, Nigeria etc. – have improved the availability of workforce stock and flow data but remain *descriptive rather than predictive*^10,11^.

To the best of our knowledge, no LMIC has developed or deployed an individual-level predictive mobility score using behavioral indicators collected through routine digital platforms. We also could not find documented evidence in literature of this practice by LMICs yet. The proposed Health Worker Mobility Score (HWMS) being proposed and piloted in Nigeria by Managedmedics.ng- a digital HR management tool for hospitals, responds to these gaps by introducing a behaviour-based, weighted scoring algorithm designed to forecast attrition and mobility risk within Nigeria’s private sector health workforce. It represents a shift from descriptive HRH monitoring to proactive, predictive intelligence aligned with contemporary global practices.

This paper presents the conceptual foundations, potential use cases, and policy relevance of the Health Worker Mobility Score within the Nigerian context. It argues that in an era of escalating health worker migration, reactive measures are no longer sufficient. What is needed is a forward-looking, data-driven mechanism that enables stakeholders: governments, hospitals, and development partners, to act ahead of the curve. By identifying those most likely to leave and supporting them early, Nigeria can not only retain critical talent but also foster a more stable and resilient health system.

## 3. Materials and Methods

### Overview

This study applies a multi-phase methodological framework to develop, evaluate, and refine the Health Worker Mobility Score (HWMS), a behavioral predictive index designed to identify early signals of internal mobility and attrition among health workers in Nigerian private hospitals. The analytical workflow integrates theory-driven indicator selection, real-world digital behavioral data, multivariable statistical modelling, machine-learning experimentation, fairness evaluation, and temporal drift monitoring. All stages adhere to the TRIPOD-AI guidance for transparent reporting of prediction models using regression and machine-learning methods, including complete specification of data sources, predictor measurement, preprocessing, modelling rationale, validation strategy, and fairness considerations.

### Conceptual and Theoretical Framework

The HWMS is grounded in labor market theory and migration theory. It recognizes that migration is rarely a one-off decision; rather, it is typically preceded by existing health labour market failures and internal mobility events, such as repeated job changes, disengagement, or skill-seeking behavior, that can serve as early warning indicators^12,13^. These signals correspond to push–pull dynamics: for example, poor remuneration and working conditions are among key factors that “push” health workers away, while overseas opportunities among others “pull” them abroad. We assume to a great extent that frequent job changes, short tenure and repeated location changes reflect discontent, whereas high participation in training, long tenure, older age may signal ongoing engagement.

The Health Worker Mobility Score^7^ (HWMS) is conceived as a predictive analytics index designed to assess and anticipate the likelihood of migration among health workers in Nigeria. It aims to transform how health facilities, digital workforce platforms, and policymakers track and respond to workforce instability in Nigeria. The score thus converts these indicators into a composite measure representing propensity for movement, consistent with human capital and expectancy theories of turnover.

### HWMS Design and Structure

The HWMS is built as a composite score derived from a set of eight (8) measurable variables linked to worker behavior, professional activity, and platform engagement. These variables are selected based on literature review and empirical evidence from a subnational HRH assessment in 2023**^Error! Bookmark not defined.^**, and are grouped into four domains:

1. Job Stability and Role History
2. Availability and Location Flexibility
3. Professional Engagement and Learning
4. Platform Behavior and Profile Completeness

As shown in Figure 1, routine workforce data (HRH logs, shifts, location) generate observable mobility events, which are transformed into a normalized, weighted Mobility Score. This score stratifies workers into risk tiers (low, medium, high), enabling targeted, supportive interventions such as planning, incentives, and behavioral support. These interventions should then influence subsequent workforce behavior, creating a continuous feedback loop.

**Figure 1:**
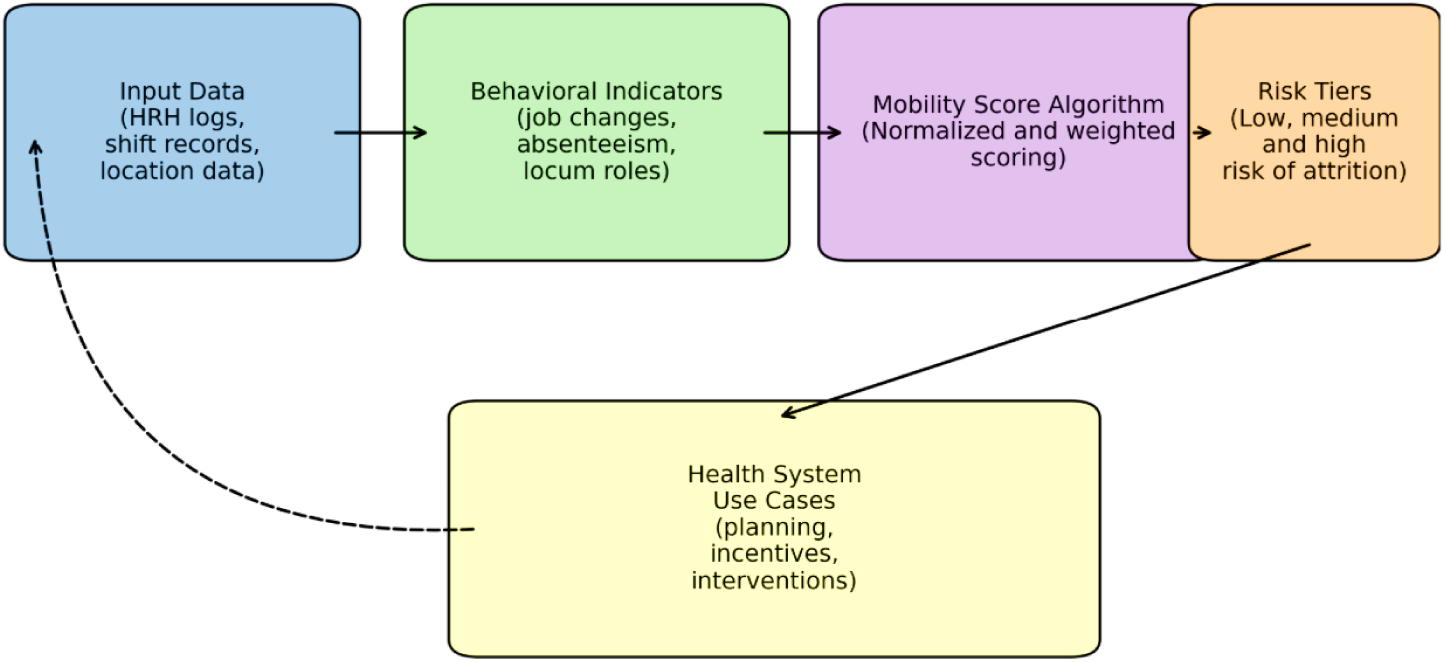
Conceptual framework for the Mobility Score.

#### Variable Measurement and Weighting

Each variable is assigned a relative weight based on its assumed predictive value, normalized between 0 and 1 for comparability, and then aggregated to generate an overall score between 0% and 100%. Table 1 below, summarizes definitions, rationales and assigned weights.

Weight assignments reflect hypothesized influence on mobility risk and will be empirically evaluated during validation and adjusted afterwards. For each worker and quarter, these variables are combined to compute a raw mobility score:

We have indicated placeholder weights to each of the core variables to guide a preliminary calculation of a health workers’ likelihood to migrate. However, to return a score that accurately predicts likelihood of migration, it is imperative that we build and train Logistic Regression models or Machine learning models that will automate this calculation using the data from managed medics, IHRIS and considerable amount of external data^8^.

Logistic Regression models like Random Forest, XGBoost, and RidgeCV are great for this purpose because they return coefficients that directly show the influence of each variable on the likelihood of migration.

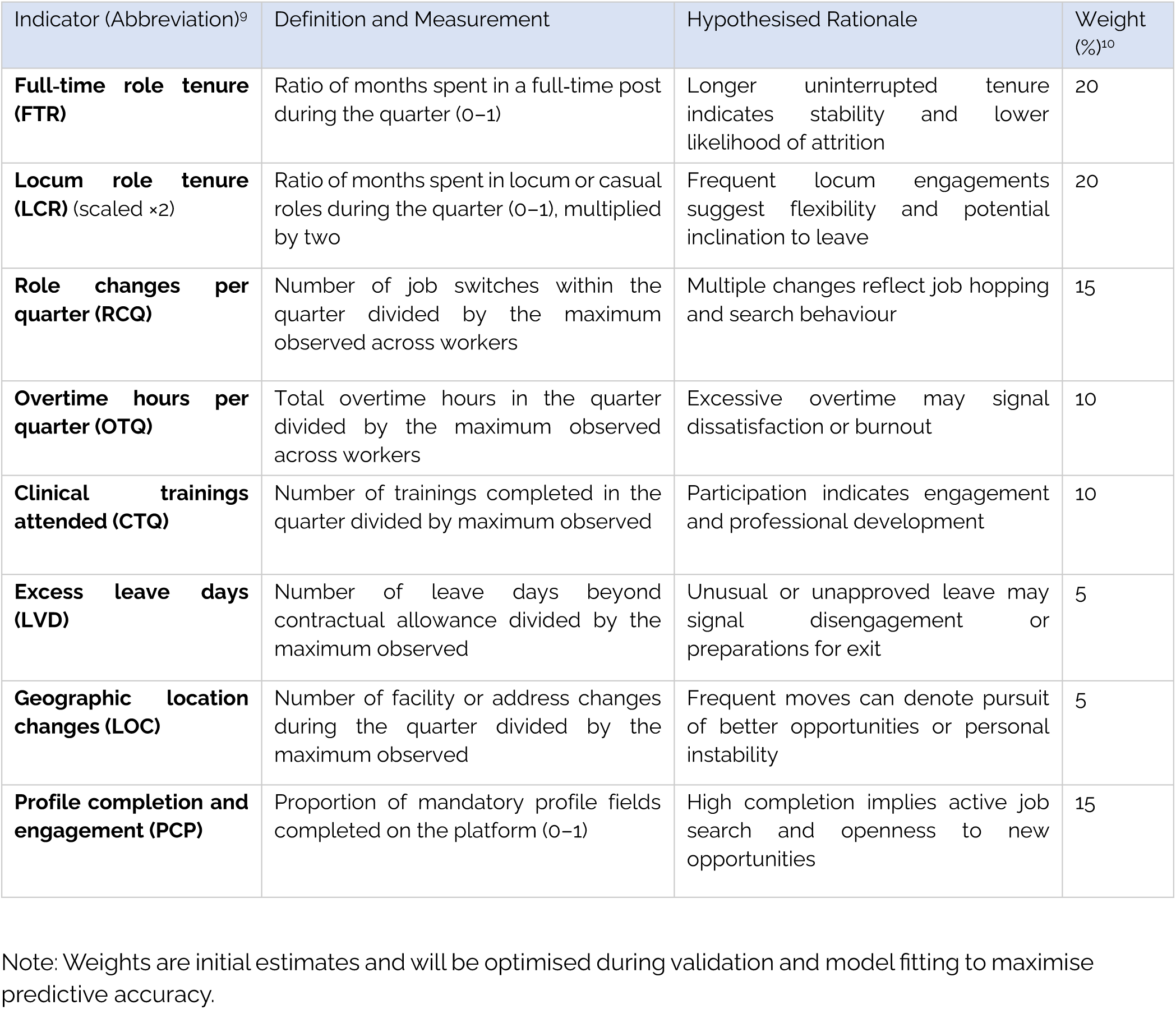

| Indicator (Abbreviation)<sup>9</sup> | Definition and Measurement | Hypothesised Rationale | Weight (%)<sup>10</sup> |
| --- | --- | --- | --- |
| **Full-time role tenure (FTR)** | Ratio of months spent in a full-time post during the quarter (0–1) | Longer uninterrupted tenure indicates stability and lower likelihood of attrition | 20 |
| **Locum role tenure (LCR) (scaled ×2)** | Ratio of months spent in locum or casual roles during the quarter (0–1), multiplied by two | Frequent locum engagements suggest flexibility and potential inclination to leave | 20 |
| **Role changes per quarter (RCQ)** | Number of job switches within the quarter divided by the maximum observed across workers | Multiple changes reflect job hopping and search behaviour | 15 |
| **Overtime hours per quarter (OTQ)** | Total overtime hours in the quarter divided by the maximum observed across workers | Excessive overtime may signal dissatisfaction or burnout | 10 |
| **Clinical trainings attended (CTQ)** | Number of trainings completed in the quarter divided by maximum observed | Participation indicates engagement and professional development | 10 |
| **Excess leave days (LVD)** | Number of leave days beyond contractual allowance divided by the maximum observed | Unusual or unapproved leave may signal disengagement or preparations for exit | 5 |
| **Geographic location changes (LOC)** | Number of facility or address changes during the quarter divided by the maximum observed | Frequent moves can denote pursuit of better opportunities or personal instability | 5 |
| **Profile completion and engagement (PCP)** | Proportion of mandatory profile fields completed on the platform (0–1) | High completion implies active job search and openness to new opportunities | 15 |

Note: Weights are initial estimates and will be optimised during validation and model fitting to maximise predictive accuracy.

#### Calculation Method (ManagedMedics Mobility Score Equation)

Let each variable be represented as follows (all normalized to a 0–1 scale):

FTR = Months in full-time role

LCR = Months in locum role (adjusted by ×2 multiplier)

RCQ = Number of role changes in a quarter

OTQ = Total overtime hours in a quarter

CTQ = Number of clinical trainings attended in a quarter

LVD = Days of leave requests above standard in a quarter

LOC = Number of location changes in a quarter

PCP = Profile completion percentage (0–1 scale)

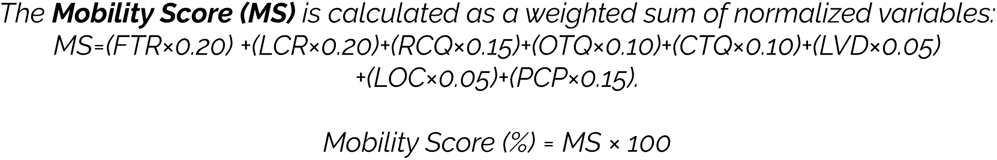

The final score ranges from 0 to 100, where higher scores indicate lower stability and higher risk of workforce mobility.

#### Risk Tiering

The final Mobility Score will allow classification of health workers into risk categories:

- Low Risk of Migration: 0–30%
- Moderate Risk of Migration: 31–60%
- High Risk of Migration: 61–100%

### Study Setting and Design

The validation study will adopt a prospective longitudinal cohort design. Health workers will be recruited from a purposive sample of private hospitals that will adopt the ManagedMedics platform, located in Lagos, FCT, Kaduna, Rivers, Imo and Ekiti. These 6 urban centres provide diversity in labour markets, patient populations and migration patterns. We will target a population of 200 hospitals across the 6 states. Hospitals will include private secondary and tertiary facilities employing doctors, nurses and midwives. Each hospital will enter a research partnership agreement specifying data-sharing protocols, roles and responsibilities.

The observation period will run for 18 months, broken into quarterly intervals. A prospective design allows temporal ordering of exposure (mobility indicators) and outcome (attrition), essential for causal inference. Multi-site recruitment enhances generalizability and permits sub-group analyses by geographic region and facility type.

### Sample Size and Power Considerations

Sample size estimation is based on detecting a meaningful association between the Mobility Score risk strata and subsequent attrition using a time-to-event framework. We assume an 18-month attrition rate of *p* = 0.25, a hazard ratio of *HR* = 1.5 between high- and low-risk groups, 80% power, and a two-sided *α* = 0.05. The required number of attrition events was calculated using the standard formula for proportional hazards models:

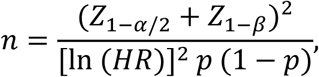

where *p*is the event proportion. Substituting the above assumptions gives an estimated requirement of approximately 175–200 events. To obtain this number of events, the minimum sample size is calculated as:

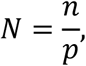

yielding approximately **700 participants**, after adjusting for a 10% loss to follow-up. To support subgroup analyses by cadre and gender, we therefore aim to recruit 800–900 health workers across all participating hospitals subject to consent and platform uptake. If attrition rates are lower than anticipated, additional hospitals may be recruited to ensure the required number of events for reliable model validation.

### Data Sourcing and Collection

#### Primary Data Collection (Predictor Data)

Primary data for the Health Worker Mobility Score (HWMS) will be obtained exclusively from the **ManagedMedics digital workforce platform**, which serves as the real-time behavioural data source for score computation. For each consenting user, the platform passively records timestamped digital activity and employment-related events, including job applications and acceptances, shift bookings, locum activity, overtime patterns, contract type (full-time vs. locum), leave requests, geographic relocation updates, credentialing actions, and profile modifications.

These platform-generated behavioural traces, linked to anonymised worker IDs: form the sole predictor dataset underpinning the HWMS algorithm. This approach ensures automated data capture, eliminates reporting burden on facilities, and provides a scalable architecture for real-time mobility risk assessment across participating hospitals. Each variable is collected over the 18-month period on the managedmedics platform and the mobility score calculates and updates quarterly. While the platform also records periods of inactivity (e.g., no login or shift activity for ≥4 months), such disengagement may reflect a range of conditions, including movement to non-partner facilities. Therefore, inactivity is treated only as a secondary signal of workforce instability, not as a primary validation outcome.

#### Secondary Data Collection (Validation Data)

Secondary data will be used exclusively for model validation and to establish robust ground-truth outcomes.

First, **administrative HR records** from participating hospitals will provide verified employment histories, including contract start and end dates, cadre, salary band, disciplinary records, reasons for exit (resignation, termination, contract lapse), and any confirmations of international migration. These HR records constitute the primary validation outcome, enabling the comparison of HWMS predictions with confirmed employment exits.

Secondly, **baseline and 6-month follow-up surveys** will be administered to all consented workers across participating hospitals to collect variables not captured in digital logs—such as socio-demographics, job satisfaction, workplace stress, remuneration perceptions, household/family factors, and intention to migrate. These data support construct validity testing, assessing whether higher mobility scores correlate with known psychosocial and motivational determinants of attrition. Together, administrative HR data and survey responses provide the external benchmarks needed to evaluate the predictive accuracy, reliability, and construct validity of the HWMS.

Cohort Inclusion criteria are: *(a)* Hospital registration and sign up on *Managed Medics*; *(b)* employment at a participating hospital at baseline; *(c)* profession as doctor, nurse, or midwife and *(d)* written informed consent for data linkage and research use. Locum staff will be included, though their shorter tenures will be accounted for in the scoring algorithm. Workers lacking reliable platform usage or declining consent will be excluded.

#### Data Preparation and Quality Assessment

Data preprocessing follows TRIPOD-AI Item 7, including:

- completeness checks and missingness mapping
- removal of inconsistent timestamps
- harmonization of facility IDs and cadre labels
- scaling and normalization of continuous predictors
- quarterly aggregation of behavioral indicators
- detection and smoothing of behavioral outliers

Missing data is handled using multiple imputation by chained equations (MICE) for survey variables, while behavioral predictors, which originate from platform activity, have minimal missingness. Workers with prolonged inactivity (>4 months) are retained but flagged for sensitivity analysis.

#### Data Analysis Plan

Data analysis will follow a structured approach combining descriptive statistics, regression modelling, survival analysis, and machine-learning techniques consistent with Transparent Reporting of a multivariable prediction model for Individual Prognosis Or Diagnosis (TRIPOD) reporting standards for predictive models.

#### Descriptive Analysis

Descriptive analyses summaries baseline characteristics, predictor distributions, and HWMS variability across time and subgroups. Longitudinal spaghetti plots and kernel density trajectories visualize temporal score patterns and behavioral drift.

#### Outcome Definitions

The **primary outcome** is **attrition**, defined as any *verified separation from employment* at a participating hospital, including voluntary resignation, contract non-renewal, or dismissal. Retirements will be excluded because they do not represent mobility-related workforce loss. Also internal transfer within the same hospital group will not count as attrition, likewise movement to non-partner facilities. (traceable only in surveys)

a) Secondary outcomes are: **confirmed international migration**, defined as documented relocation abroad for employment, licensing, or training;
b) **time to attrition**, defined as the interval from baseline to the first verified separation event.

#### Construct Validation

Construct validity tests whether the HWMS behaves in theoretically expected ways. Mixed-effects models examine correlations between mobility scores and psychosocial constructs such as job satisfaction, burnout, and migration intention. We expect HWMS to correlate negatively with satisfaction and positively with migration intent, consistent with established turnover determinants^14^ ^15^ ^16^. Between-group comparisons assess whether cadres, gender groups, or regions exhibit systematically different score distributions in line with contextual evidence.

### Predictive Modelling (Regression-Based Approaches)

#### Logistic Regression (Binary Attrition)

A multivariable logistic regression model predicts overall attrition. Predictors include the HWMS, demographic covariates (age, gender, cadre), facility type, and state. Model performance is evaluated using:

- AUC (discrimination)
- Brier score
- calibration slope and intercept
- Hosmer–Lemeshow goodness-of-fit test
- calibration plots using flexible splines

To mitigate class imbalance, we apply techniques such as class-weighted loss functions or SMOTE variants where appropriate.

#### Cox Proportional Hazards (Time-to-Exit)

A Cox model examines time to attrition, incorporating the same predictors. Proportional hazards assumptions are tested using Schoenfeld residuals and time-interaction terms. Baseline hazard functions and survival curves illustrate temporal risk profiles.

#### Machine Learning Modelling

To test whether flexible models outperform regression, we develop Random Forests, XGBoost, Support Vector Machines, and feed-forward neural networks. Model development follows TRIPOD-AI Items 12c–12e, including:

- hyperparameter tuning via 10-fold cross-validation
- optimization using grid search and Bayesian optimization
- documentation of all hyperparameters
- reporting of feature importance using SHAP values

Performance metrics include AUC, PR-AUC, Brier score, sensitivity, specificity, and calibration curves. A model is eligible for adoption only if it improves AUC by ≥0.05 over logistic regression or shows substantially better calibration.

#### Fairness and Subgroup Analysis

Fairness evaluation examines model performance across gender, cadre, salary bands, and regions. Metrics include:

- subgroup-specific AUC
- calibration parity (slope deviation ≤ ±0.10)
- error symmetry (balanced false-positive/false-negative rates)

Mitigation strategies include subgroup-specific calibration, threshold adjustment, and reweighting.

#### Error Analysis

To ensure model safety, we examine patterns of misclassification:

- false-positive and false-negative distributions
- subgroup-specific error burdens
- qualitative interpretation of misclassified cases

This phase informs threshold optimization and harm mitigation.

#### Temporal Validation and Drift Detection

Each quarter, we assess:

- **data drift** (predictor distribution shifts)
- **behavioral drift** (changes in worker activity patterns)
- **calibration drift**
- **predictive drift** (AUC decline)

If AUC declines by >0.05 or calibration slope deviates by >0.15, models undergo recalibration or full retraining.

#### Sensitivity Analyses

Sensitivity analyses include:

- alternative attrition definitions
- exclusion of inactive workers
- alternative score weighting schemes
- tercile vs quartile cutoffs
- robustness of predictors across cadres

These analyses test the stability of model conclusions.

#### Transparency, Reproducibility, and Open Science

In line with TRIPOD-AI Items 18a–18f, all model equations, code, hyperparameters, calibration plots, and fairness evaluations will be documented. Where permissible, anonymized datasets and analytical code will be shared under controlled-access conditions.

### VALIDATION FRAMEWORK AND MODEL TESTING PLAN

#### Purpose of Validation

The validation framework ensures that the HWMS prediction model is accurate, generalizable, fair, temporally stable, and safe for operational use in HR workflows. The process integrates statistical, technical, ethical, and human-centered validation layers, consistent with TRIPOD-AI recommendations for evaluating prediction models that use regression or machine-learning approaches.

The objectives of validation are to:

1. Evaluate predictive discrimination and calibration.
2. Assess generalizability across hospitals, states, and worker subgroups.
3. Ensure fairness across demographic and professional groups.
4. Detect prediction drift over time.
5. Ensure usability, interpretability, and safety within HR operations.

#### Internal Validation

Internal validation constitutes the first stage of evaluation. The development dataset is partitioned using a 70/30 train–test split, complemented by 10-fold cross-validation and extensive bootstrapping to estimate optimism and guard against overfitting. Performance is quantified through area under the ROC curve (AUC), calibration slope and intercept, Brier scores, and graphical calibration assessments.

Models must demonstrate an AUC of at least 0.70, good alignment between predicted and observed risks, and minimal discrepancy between apparent and validated performance. Decision curve analysis is additionally used to determine whether the model confers net benefit across clinically meaningful risk thresholds.

#### Cross-Site Quasi-External Validation

Because all participating facilities operate on the Managed Medics platform, a quasi-external validation strategy is employed to approximate independent evaluation. This is achieved through Leave-One-Hospital-Out (LOHO) and Leave-One-State-Out (LOSO) procedures, which test the model’s stability when exposed to unseen organizational cultures, labor markets, and workforce characteristics.

A model that performs robustly across these withheld clusters is considered generalizable within the private-sector ecosystem. In cases where performance deteriorates markedly, remedial actions such as recalibration, feature simplification, or the incorporation of facility-level random effects are considered.

#### Fairness Evaluation

Ensuring fairness across worker subgroups is critical for ethical deployment. Model performance is examined separately for cadres, genders, regions, salary bands, and facility types. Subgroup-specific discrimination, calibration, and error rate symmetry are evaluated in accordance with TRIPOD-AI guidance on fairness reporting.

A model is considered equitable when calibration slopes across subgroups remain close to unity and performance metrics do not diverge meaningfully. Should any subgroup exhibit systematic disadvantage, such as disproportionately high false-positive or false-negative rates, mitigation strategies, including subgroup-specific recalibration or threshold adjustments, are applied to restore equity^17^.

#### Temporal Validation and Drift Monitoring

Given the dynamic nature of Nigeria’s labor market, temporal validation forms a core layer of the evaluation process. The HWMS model is monitored quarterly to detect data drift (changes in predictor distributions), behavioral drift (shifts in worker mobility patterns), calibration drift, and predictive drift. Trends are assessed by comparing successive quarters of model performance to the original internal validation benchmarks.

Minor drift may be resolved through recalibration techniques such as Platt scaling or isotonic regression, while substantial performance degradation triggers full retraining of the model followed by renewed internal and quasi-external validation. This ensures the model remains relevant as workforce conditions evolve.

#### Human-Centered and Ethical Validation

Validation extends beyond statistical criteria to include an assessment of how end-users understand and interact with the model. Employers evaluate the performance of their HR Managers based on how well they interpret the output of SHAP (SHapley Additive explanations) and Risk Outputs and the performance of HR Managers as indicated by their evaluations of polygons (calibration plots) or through structured interviews, as well as through surveys to assess worker perceptions of Fairness, Transparency, and Psychological Effects.

Potential risks associated with the misunderstanding or misuse of the results of Scenario-Based Safety Testing may include punitive reactions from supervisors toward employees’ Risk Scores that are considered elevated or the reinforcement of existing workplace inequities. To minimize these risks, Governance Safeguards have been implemented, including, among others, requiring HR Managers to supervise the use of Human Resources (HR) Models, requiring HR Managers to provide a Justification for any HR Decision that is influenced by the Results of a Model, and conducting regular Ethical Audits to ensure continued Compliance with Responsible AI Principles^18^ ^19^.

#### External Validation Roadmap

Although external validation lies beyond the scope of the current study phase, a forward plan is established to evaluate the model across public hospitals, additional geographic regions, and new worker cohorts over time. These future evaluations will apply the same discrimination and calibration benchmarks used in the internal and quasi-external validations, ensuring continuity and comparability across settings.

#### Model Testing Plan

Multiple algorithmic approaches are evaluated to determine the optimal modelling strategy. Logistic regression and Cox proportional hazards models serve as interpretable baselines, while Random Forests, XGBoost, Support Vector Machines, and neural networks are explored to determine whether more flexible models offer meaningful improvements. All models undergo a uniform development pipeline consisting of preprocessing, hyperparameter optimization, cross-validated performance estimation, fairness testing, and interpretability assessment using SHAP values. Performance is compared using AUC, precision–recall AUC, Brier scores, sensitivity, specificity, and calibration quality.

#### Threshold Selection and Risk Tiering

The risk of placing workers into their respective risk categories (low, medium and high) is based upon a combination of statistical optimization (using the Youden index as an example), decision curve analysis, and analyzing how the classification system will impact on false classifications. The appropriateness of the thresholds is evaluated in subgroups to ensure that they do not disproportionately burden specific groups of workers. In cases where thresholds require adjustment, subgroup-specific thresholds or recalibration strategies will be employed to ensure equitable treatment of all workers.

#### Failure Mode and Harm Analysis

A methodical examination of failure modes identifies all of the possible conditions that could create adverse outcomes due to model predictions. These include evaluating false positive (incorrect positive) and false negative (incorrect negative) rates, identifying which groups of customers have the highest chance of being incorrectly identified, and evaluating how these errors can affect negative HR patterns. As a response to identified failures, corrective actions, such as creating alternative thresholds or providing greater oversight from humans when making decisions about high-risk candidates, have been included in the governing plan.

#### Deployment Readiness Criteria

A model is only considered ready for deployment in a live environment when it shows high levels of discrimination, good calibration, equality across demographic subgroups, consistent performance over multiple datasets over time, clearly interpretable through SHAP-based explained values, and minimal risk during operation. If there are multiple models that have passed these criteria, the preference will be given to the simplest and most interpretable model, as indicated by the emphasis put on both transparency and utility by TRIPOD-AI.

#### Post-Deployment Monitoring

Model performance is continually monitored on a quarterly basis, post-deployment, with regard to all standard metrics that indicate potential issues with discrimination, calibration, fairness, and drift. On an annual basis, the HWMS model undergoes an ethics and fairness audit that evaluates adherence to the responsible use of AI principles, and user feedback is incorporated from HR, in order to improve workflow and enhance interpretability of results generated by the HWMS model. The HWMS model is re-trained at least once every 6-12 months; however, if observed drifts exceed acceptable levels, then retraining will happen as needed. All of these ongoing checks & balances provide the assurance that the HWMS Model will reliably perform ethically and fairly, both in the short term and long term.

## 4. Results (Conceptual Illustration)

The results section, is entirely conceptual and presents the output of the HWMS computation after implementation. To illustrate the HWMS calculation, consider a hypothetical worker who spends the entire quarter in a full-time role (FTR = 1.0), completes no locum shifts (LCR = 0.0), changes roles twice (RCQ = 0.67), works four hours of overtime (OTQ = 0.33), attends one training session (CTQ = 0.33), takes two excess leave days (LVD = 0.28), changes location once (LOC = 0.33) and completes 90 % of their profile (PCP = 0.90). Plugging these values into the formula yields a Mobility Score of 53.2 %, classifying the worker as moderate risk. While purely hypothetical, this example demonstrates how diverse behavioural indicators translate into a risk tier and highlights the need for empirical calibration.

| Variable | Normalized Value | Weight<sup>11</sup> | Weighted Score |
| --- | --- | --- | --- |
| Full-time role (3 of 3 months) | 1.0 | 20% | 0.20 |
| Role changes (2 of 3) | 0.67 | 15% | 0.10 |
| Overtime (4 of 12 hours) | 0.33 | 10% | 0.033 |

<sup>11</sup> Weights presented are placeholders. Final weights will be fully data-driven following model training.

|  |  |  |  |
| --- | --- | --- | --- |
| Clinical training (1 of 3) | 0.33 | 10% | 0.033 |
| Leave (2 of 7 days) | 0.28 | 5% | 0.014 |
| Location changes (1 of 3) | 0.33 | 5% | 0.017 |
| Profile completion (90%) | 0.90 | 15% | 0.135 |

Total Mobility Score = 0.532 (or 53.2%) for the quarter

### Synthetic Mobility Score Distribution (Histogram)

**Figure 2:**
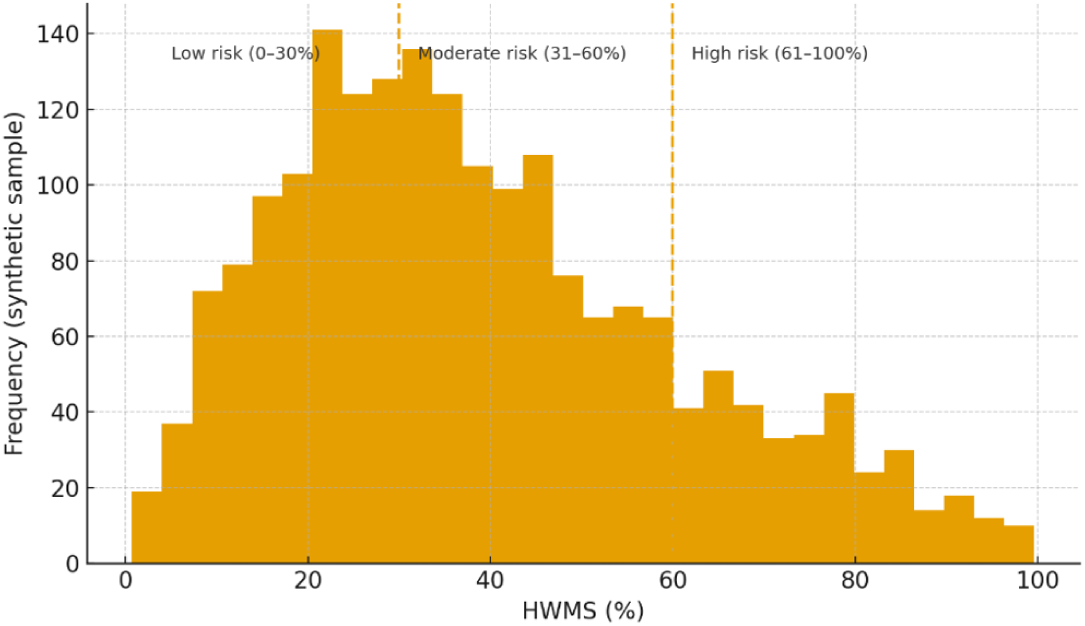
Synthetic Distribution of the Health Worker Mobility Score (HWMS) illustrating expected clustering across low-, moderate-, and high-risk tiers (0–30%, 31–60%, 61–100%). This conceptual pattern reflects how behavioral indicators may cluster across mobility risk tiers.

Figure 3 presents a synthetic distribution of HWMS values, illustrating the expected scoring pattern and the proportion of workers falling into low-, moderate-, and high-risk categories. This conceptual histogram provides an early visual intuition for how behavioral indicators may aggregate across mobility risk tiers.

**Figure 3:**
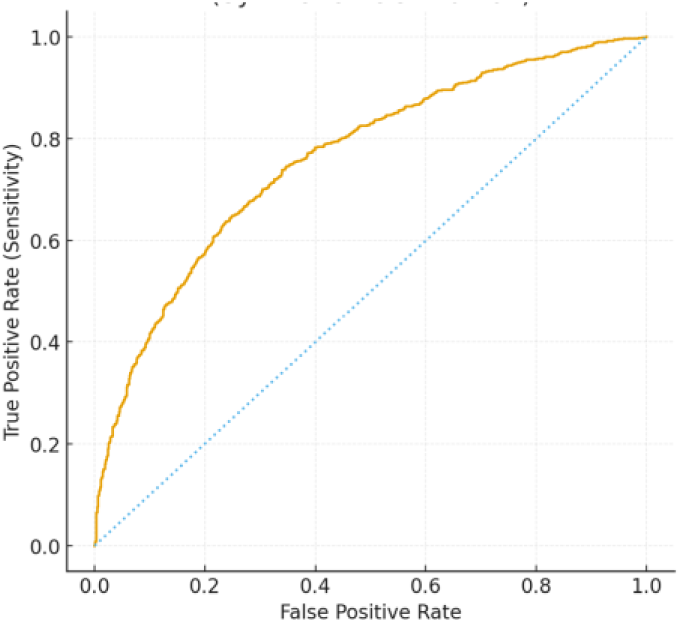
Conceptual receiver operating characteristic (ROC) curve for the Health Worker Mobility Score (HWMS). The curve is generated from synthetic data to illustrate expected discriminative performance; the synthetic AUC (0.761) is shown for illustrative purposes only. Final AUC values will be reported after empirical validation.

### Conceptual ROC Curve for the HWMS (Expected Predictive Performance)

Figure 4 shows a conceptual ROC curve for the HWMS with a synthetic AUC of 0.761; this illustration demonstrates the type of discriminative performance we may expect to evaluate during model validation.

**Figure 4:**
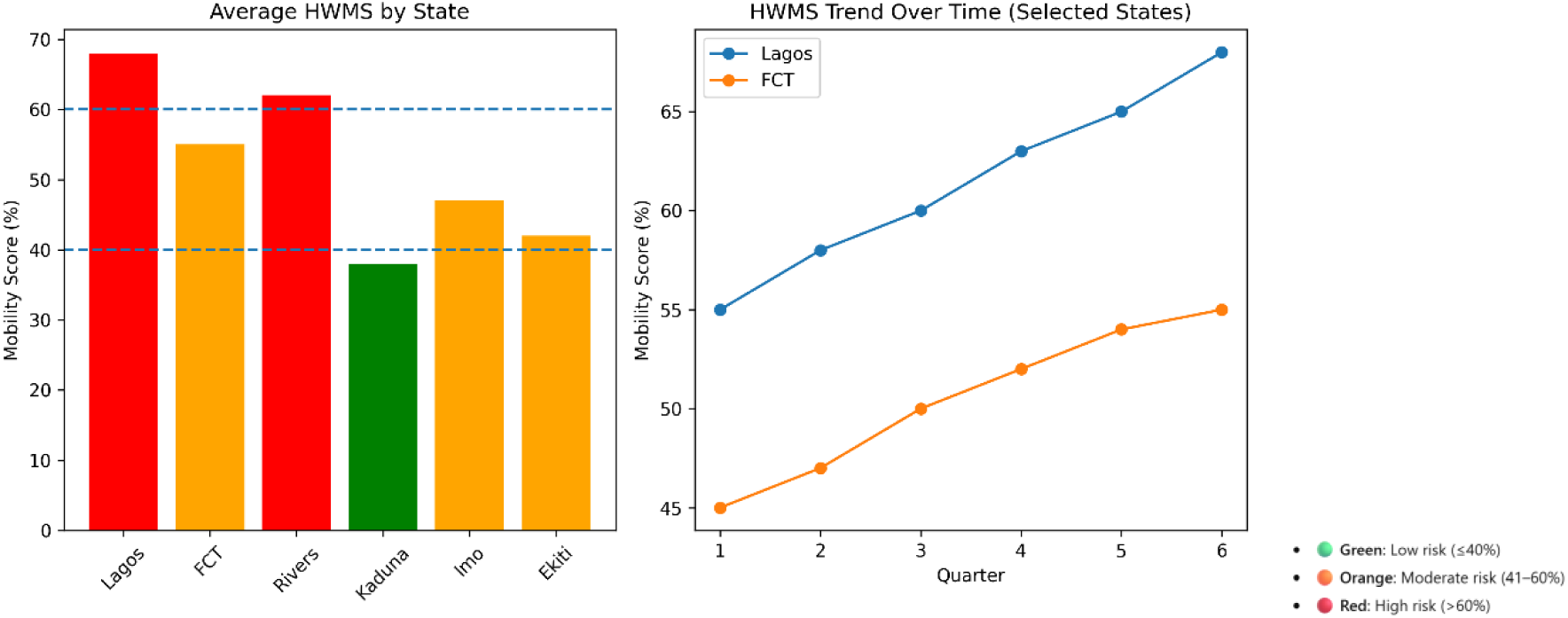
Conceptual state level HWMS Decision support visualization- At the macro level, aggregated HWMS outputs can be visualized as color-coded state dashboards, displaying average mobility risk alongside longitudinal trend graphs. Such visualizations enable policymakers to rapidly identify high-risk regions, monitor deterioration or improvement over time, and priorities targeted retention or managed migration interventions

## 5. Discussion

### a) Key Contributions and Interpretation

This study introduces the Health Worker Mobility Score (HWMS) as a behaviour-based, real-time approach to identifying attrition and migration risk within Nigeria’s health workforce. Traditional HRH planning tools in Nigeria rely heavily on retrospective administrative headcounts and health workforce-to-population ratios, which do not capture the dynamic behavioural signals that precede workforce exit. By integrating indicators such as job changes, locum intensity, absenteeism, training participation and digital engagement patterns, the HWMS reframes workforce instability as a *measurable*, *predictable* continuum rather than a sudden event.

The conceptualization aligns with push–pull migration theory^20^ and capital theory, whereby economic hardship, insecurity and poor job satisfaction manifest early as job-search behaviour, locum shifts or platform disengagement. The score also operationalizes labour-market segmentation theory by detecting transitions from lower-security to higher-security labour segments, including international migration pathways. In this way, the HWMS translates behavioral micro-signals into actionable risk categories, offering an innovative mechanism for proactive HRH management in Nigeria’s rapidly evolving private health sector.

### b) Alignment with Global and National Frameworks

The design and proposed use of the HWMS align strongly with global digital-health and workforce governance frameworks. The WHO Global Strategy on Digital Health (2020–2025) underscores the need for integrated data systems and predictive analytics to strengthen UHC and support resilient health systems. Similarly, Nigeria’s National Policy on Health Workforce Migration (2023) calls for improved retention mechanisms, ethical recruitment, strengthened HRH information systems and real-time labour-market intelligence to support the country’s shift toward *managed migration*.

By generating early-warning signals, the HWMS directly supports these policy imperatives. Aggregated risk data can inform implementation of in-service training reforms, help identify occupational groups (including their seniority and experience levels) vulnerable to international recruitment pressure, and guide national and state-level planning for deployment, recruitment and investment in working conditions. Furthermore, integrating the score into systems such as Nigeria’s iHRIS platform or the ManagedMedics ecosystem offers a practical pathway to achieving interoperability between private-sector digital health tools and government HRH systems: a documented gap highlighted in national HRH assessments.

Importantly, the HWMS offers a structured mechanism for operationalising Nigeria’s managed migration policy. By identifying where migration is safe or risky, the score can guide negotiations with destination countries, support ethical recruitment agreements and determine where retention incentives or safeguards are required.

### c) Comparisons with Existing Models and Novelty

Predictive workforce analytics are increasingly utilised in high-income settings, where hospitals deploy machine-learning models to anticipate nurse turnover using structured payroll data, performance metrics and attendance records. However, these models often operate in environments with mature HR information systems and formal labour markets. Nigeria’s health workforce landscape—characterised by fluid movement between formal and informal roles, widespread locum practices, inconsistent record keeping and rising international recruitment, requires contextual adaptations.

The HWMS is novel in several respects. First, it incorporates behavioural and labour-market signals that are peculiar to LMICs, such as high locum intensity, credential verification patterns and digital engagement proxies. Second, it offers transparency in indicator selection and weighting, in contrast to “black-box” proprietary algorithms. Third, the design includes ethical and governance safeguards tailored to Nigeria’s regulatory and labour context, ensuring that predictive analytics are used for supportive retention rather than punitive measures. Through its public conceptualization and multi-stakeholder oversight, the HWMS contributes to responsible AI development in African health systems.

### d) Policy and Programmatic Implications

The HWMS has multilevel implications for Nigeria’s health sector. At the facility level, the score supports targeted retention interventions, including training opportunities, mentorship, financial incentives, flexible scheduling and well-being support, directed toward individuals at the highest risk of attrition. Low-risk individuals can be identified for leadership development, improving career trajectories and strengthening institutional culture.

At the state level, aggregated mobility risk maps can help health commissioners anticipate shortages, refine deployment strategies and calibrate recruitment pipelines. State HRH units can use quarterly mobility trends to forecast staffing needs, detect stress points in the labour market and design tailored retention packages for critical cadres.

At the national level, the score provides a critical evidence layer for workforce planning, compensation reforms, ethical recruitment strategies and the implementation of Nigeria’s managed migration policy. The HWMS can inform bilateral agreements with destination countries by identifying occupational groups where outward migration is feasible and those where further losses would threaten service continuity. By integrating the score into national HRH registries, policymakers can monitor progress toward retention targets, assess distributional equity and align investments with Nigeria’s UHC roadmap.

### e) Theoretical and Scientific Contributions

Beyond operational utility, the HWMS advances theoretical debates on workforce mobility and migration by demonstrating how micro-level behavioural traces can predict macro-level labour outcomes. It provides an empirical platform for testing hypotheses about the sequence of events leading from dissatisfaction and job-search behaviour to domestic job changes and ultimately international migration. The longitudinal structure of the score also enables exploration of mobility trajectories over time, enriching scholarship on migration as a process rather than an event.

Methodologically, the study contributes to the validation literature for predictive analytics in LMIC settings by combining platform-based behavioural data, administrative HR records and worker-reported motivations—an integrated approach rarely attempted in African HRH research. Findings may inform similar tools in other critical sectors such as education, policing and public administration.

#### Proposed Use cases of the HWMS

I. **Predictive analytics:** Real-time monitoring of mobility scores will flag individuals or units at risk of attrition, enabling HR managers to initiate conversations and support mechanisms before formal resignations occur. Such early warning systems are common in corporate HR analytics and can be adapted to health care settings facing workforce volatility.
II. **Targeted retention strategies:** High-risk workers may be prioritized for financial incentives, career development programmes, mentorship, housing allowances or flexible contracts, while low-risk workers could be groomed for leadership roles. By tailoring incentives to risk profiles, hospitals can allocate resources efficiently and improve retention outcomes.
III. **Strengthened Workforce planning:** Aggregated mobility scores across facilities and regions can inform state and national planners about impending staffing gaps. Data visualizations of risk hotspots can aid in deploying experienced personnel to underserved areas, adjusting recruitment pipelines and forecasting training needs.
IV. **Embedded Decision Support Tool:** Assuming potential collaboration from the Federal Ministry of Health and the medical regulatory bodies, the HWMS predictive analytics can be embedded into national HR information systems—such as iHRIS, to create a unified metric for workforce risk. In this context, the score can be shown in multiple ways. Firstly, as shown in figure 4 below, HWMS Predictive Analytics can be color-coded (based on risk score) and placed adjacent to Trend Graphs that show the evolution of Employees Mobility Score over time. Secondly, HWMS Predictive Analytics can include an explanation Panel that provides transparency regarding the factors that drove the prediction. This supports the early-warning and data-governance objectives outlined in Nigeria’s National Policy on Health Workforce Migration.
V. **Managed migration:** The mobility score can serve as a practical instrument for operationalizing Nigeria’s workforce migration policy, enabling government to identify occupational groups or regions where outward migration may be ethically permissible. By stratifying facilities or states by migration-risk levels, the HWMS can help policymakers determine: (a) where bilateral agreements with destination countries can be supported; (b) where additional safeguards or retention investments are required; and (c) how to align migration pathways with domestic staffing thresholds. In this way, the score provides an objective, data-driven basis for implementing the country’s migration policy while protecting service continuity in vulnerable areas.

## 6. Limitations and Challenges

This study faces several limitations.

I. First, platform-based behavioural data may vary in completeness depending on facility adoption and user engagement. Workers who do not consistently use ManagedMedics may generate sparse behavioural traces, potentially reducing predictive accuracy.
II. Second, the score may not fully capture socio-cultural drivers of migration; such as family pressures, social networks abroad or macroeconomic shocks, which remain difficult to quantify.
III. Third, algorithmic bias is a risk if demographic groups systematically exhibit behaviours that map onto higher risk scores. This requires deliberate fairness audits, demographic adjustment and stakeholder oversight.
IV. Fourth, the study focuses on private-sector hospitals; generalising findings to public facilities—which operate under different employment conditions—will require further validation.
V. Finally, there is potential for misuse: without strong governance, mobility scores could be interpreted punitively (e.g., to deny promotions or penalise staff). This underscores the necessity of a robust ethical framework, transparent communication and clear policy guidance.

### 7. Conclusion and Future Directions

Nigeria’s health workforce crisis is no longer defined solely by shortages, but by instability driven by continuous internal mobility and accelerating international migration. Existing HRH information systems, largely retrospective and descriptive are structurally incapable of detecting the behavioral signals that precede workforce exit. This paper responds to that gap by conceptualizing the Health Worker Mobility Score (HWMS): a behavior-based, real-time predictive analytics construct designed to identify early warning signs of attrition within Nigeria’s health workforce.

The HWMS advances digital health practice in three important ways. First, it reframes health worker migration as a predictable process rather than a sudden event, translating micro-level behavioral signals, such as job switching, locum intensity, absenteeism, training engagement, and platform activity into actionable risk stratification. Second, it demonstrates how routinely generated digital workforce data can be repurposed into early-warning intelligence, enabling proactive and supportive retention strategies at facility, state, and national levels. Third, it embeds predictive analytics within a governance-aware and ethically grounded framework, explicitly addressing fairness, interpretability, drift monitoring, and safeguards against punitive misuse—concerns that remain underdeveloped in many digital HR and AI deployments.

From a systems perspective, the HWMS offers a practical mechanism for operationalizing Nigeria’s policy shift toward managed health worker migration. Aggregated mobility risk patterns can inform workforce planning, guide retention investments, support ethical recruitment negotiations, and strengthen interoperability between private-sector digital platforms and national HRH registries such as iHRIS. Importantly, the score is designed not as an automated decision-making tool, but as decision support, augmenting human judgment rather than replacing it.

This study is intentionally forward-looking. The current contribution lies in the conceptual design, theoretical grounding, and validation framework of the HWMS rather than in reported empirical performance. The proposed prospective longitudinal cohort study will be critical to empirically testing predictive accuracy, calibrating indicator weights, and assessing feasibility for scale-up. Future work should also examine how health workers perceive algorithmic risk assessment, how managers interpret and act on mobility signals, and whether interventions triggered by the score measurably improve retention outcomes.

In an era of intensifying global competition for health talent, LMICs require tools that move beyond counting health workers toward anticipating loss and enabling early action. The Health Worker Mobility Score represents a step in that direction: demonstrating how digital health systems can generate predictive, ethically governed workforce intelligence to support more resilient health systems.

## Data Availability

This manuscript does not report analyses of empirical data. It presents the conceptual design and proposed validation framework of the Health Worker Mobility Score. No datasets were generated or analyzed during the current study. All data to be collected during the proposed prospective longitudinal validation study—including de-identified platform-generated behavioural data, administrative human resources records, and survey responses—will be made available in accordance with the journal Data Policy upon completion of the study, subject to ethical approval and participant consent. De-identified datasets and accompanying metadata will be deposited in a publicly accessible data repository, or made available upon reasonable request if restrictions are required to protect participant privacy and confidentiality

## 8. Declarations

- **Conflicts of Interest**: The authors declare that they have no competing interests.
- **Author Contributions:**

- **Chukwuemeka Azubuike** conceptualized the study, led the development of the Health Worker Mobility Score framework, and designed the overall methodological and validation approach. He drafted the original manuscript, coordinated revisions, and takes responsibility for the integrity of the work as a whole.
- **Yusuf Ajiboye** contributed to the methodological design, particularly the statistical modelling, validation framework, and analytic strategy. He supported the development of the predictive modelling plan and reviewed the manuscript for methodological accuracy.
- **Onyema Ajuebor** provided strategic and policy-level input, contributed to the interpretation of findings within global health workforce and migration frameworks, and reviewed the manuscript for alignment with international health systems and governance perspectives.
- **Oyebanji Filani** contributed to the contextualization of the study within Nigeria’s health system, supported interpretation of programmatic and implementation implications, and reviewed the manuscript for policy relevance and clarity.
- All authors reviewed and approved the final manuscript.
- **AI Use Disclosure**: The authors affirm that this manuscript was conceived, analysed, and written by the authors. Artificial intelligence tools (specifically ChatGPT, Perplexity) were used solely for language refinement, grammar correction, and sentence formatting suggestions during the drafting process. No AI tools were used to generate or analyze data, and all intellectual contributions, including conceptual framing and writing decisions, remain the authors’ sole responsibility.

## Footnotes

5 Nigeria’s health workforce faces challenges which play out along a spectrum of health worker production, deployment and distribution, retention, and mobility. The spectrum in a well-functioning health system should facilitate a seamless flow of health workers along this continuum and minimize excessive turnover and brain drain by addressing the root causes of dissatisfaction and provide incentives for health workers to remain within the system.

6 Based on KIIs with HRH stakeholders during roundtable discussions in 2023 across the 6 geopolitical zones in Nigeria.

7 The ManagedMedics platform, enables a dynamic calculation and continuous updates of the Mobility Score based on real-time data collection.

8 Typically, 2000 rows for regression data and 5000 – 1000,000 rows of data for ML models.

9 The current conceptual framework does not explicitly incorporate comparative financial earnings or wage differentials, despite strong evidence that income disparities are a key driver of health worker mobility and international migration. This omission reflects both data availability constraints and a deliberate design choice to prioritize behavioral indicators that can be passively and consistently captured through digital workforce platforms. As a result, the mobility score may under-estimate risk among workers whose primary motivation for movement is financial rather than behavioral. Future iterations of the Health Worker Mobility Score will explore the inclusion of relative earnings indicators—such as deviations from cadre- and location-specific pay benchmarks or ratios of locum to base earnings—subject to data governance safeguards and empirical validation.

10 Weights presented are placeholders. Final weights will be fully data-driven following model training.

11 Weights presented are placeholders. Final weights will be fully data-driven following model training.

